# Global Disparities in Vaccine Reimbursement for Adults Living with HIV: A Cross-Sectional Survey of 120 Countries by EVASG and ESTISG

**DOI:** 10.64898/2026.10.30.26366946

**Authors:** Emily Burzynski, Özlem Türkmen Recen, Petar Velikov, Ahmed Cordie, Gabriela Zambrano, Alfonso J. Rodriguez-Morales, Manuel Krone, Olivier Epaulard, Zitta Barrella Harboe, Justyna Kowalska

## Abstract

**Objective:** Adults living with HIV are at increased risk of vaccine-preventable infections, yet access to recommended vaccines varies globally. We evaluated national vaccination guidelines, public reimbursement, and on-site vaccination availability across countries with different income levels.

**Methods:** We conducted a cross-sectional survey of HIV clinicians and experts representing 120 countries. The 26-item questionnaire assessed vaccination guidelines, recommendations, public reimbursement, and on-site vaccination availability. Countries were classified according to World Bank income group. Generalized linear mixed-effects models assessed routine recommendations, full reimbursement, and recommendation-reimbursement gaps by vaccine type and income group.

**Results:** A total of 124 responses representing 120 countries were analyzed, including 38 low-/lower-middle-income (LIC/LMIC), 36 upper-middle-income (UMIC), and 50 high-income countries (HIC). HIV-specific national vaccination guidelines were reported by 37% of respondents, increasing from 18% in LIC/LMIC to 52% in HIC (*p*=0.026). Only 30% reported full public reimbursement for all recommended vaccines (13% LIC/LMIC, 28% UMIC, 44% HIC;*p*=0.007). Mixed-effects analyses showed significant differences in routine recommendations and full reimbursement by vaccine type and income group. The adjusted probability of routine recommendation was lower in LIC/LMIC than HIC countries (52.6% vs 77.6%; adjusted p=0.0013). Among recommended vaccines, more than 94% of influenza, pneumococcal, and hepatitis B vaccinations lacked full reimbursement in LIC/LMIC, based on adjusted estimates. Reimbursement gaps for HPV and herpes zoster vaccines remained substantial even in HIC. On-site vaccination was also less available in LIC/LMIC than in HIC (55% vs 78%; p=0.020).

**Conclusions:** Significant income-related disparities persist in vaccination policy and implementation for adults living with HIV. Gaps between recommendations and financial access are particularly pronounced in lower-income settings, while HPV and herpes zoster reimbursement remains limited across all income groups. Expanding HIV-specific vaccination policies, improving public reimbursement, and integrating vaccination into routine HIV services are essential to improve access to recommended vaccines for this population.

## Background

Adults living with HIV are at increased risk of vaccine-preventable diseases, partly due to persistent immune dysfunction that may impair humoral and cellular immune responses to vaccination [1,2]. International guidelines provide recommendations on vaccination for adults living with HIV; however, their translation into national policies and implementation across different healthcare settings has not been globally studied [1,3].

Public reimbursement strategies vary globally and depend on local health priorities, while vaccine market access is often jeopardized by geography, sales politics, and social prejudices [4,5]. These factors may influence whether recommended vaccines are accessible, particularly in relation to country’s income level. Financial and structural barriers may differ across income groups, with vaccine procurement and healthcare financing influencing access in both lower and higher income settings [6,7]. Conversely, high-income countries might struggle with brand product prices irrespective of well-functioning systems and benefit awareness. In result, suboptimal vaccine uptake has also been reported in high-income countries [8,9], showing that barriers to vaccination are a global issue and extend beyond simple differences in national income. Global data comparing reimbursement of recommended vaccines for adults living with HIV remain limited.

In this study, we aimed to assess vaccination recommendations and public reimbursement for adults living with HIV across countries of different income levels and to identify gaps between vaccine recommendation and financial access.

## Methods

We conducted a cross-sectional survey assessing HIV specific vaccination guidelines, public reimbursement policy, and availability of vaccination for adults living with HIV globally. One expert per country, familiar with national vaccination recommendations, reimbursement policies, and practical implementation, was invited to participate. Participants were identified through professional networks of the Euroguidelines in Central and Eastern Europe (ECEE) Network Group, ESCMID Vaccine Study Group (EVASG) and ESCMID Sexually Transmitted Infections Study Group (ESTISG), with regional contacts supporting recruitment in Latin America, Africa, and the Middle East.

The 26-item questionnaire (23 closed- and three open-ended questions) was designed by EVASG Executive Committee members. Seven vaccines were prioritized for evaluation: influenza, pneumococcal, hepatitis B, tetanus-diphtheria-acellular pertussis (Tdap), measles-mumps-rubella (MMR), human papillomavirus (HPV), and herpes zoster (VZV). The questionnaire assessed the following fields: respondent characteristics, HIV-specific vaccination guidelines availability by country, the specific recommendations for seven selected vaccines, reimbursement strategy, on-site vaccination availability, and barriers to provision of vaccines to people living with HIV (Supplementary Material S1).

Paper questionnaires were first distributed at the ECEE Network Group Conference in Warsaw, Poland on 16 June 2025. Following EVASG and ESTISG Executive Committee approval, an identical electronic questionnaire was distributed through both study groups in November 2025. Participants were informed of the purpose of the study, and participation was voluntary, with completion of the questionnaire indicating agreement to participate. Where vaccination or reimbursement policies varied within a country, responses from individual regions or provinces were included separately to reflect these differences. For the analyses, each country or separately represented subnational region or province was considered one jurisdiction.

Countries were grouped according to the 2025 World Bank income classification as low- and lower-middle-income (LIC/LMIC), upper-middle-income (UMIC), or high-income (HIC).

Self-reported vaccine recommendation practices were classified as routine, conditional/selective, or not recommended/actively promoted. Routine recommendations included consistent vaccine promotion, whereas conditional/selective recommendations included those restricted to specific groups, clinical circumstances, or inconsistently provided because of time constraints. The primary outcome compared routine recommendation with conditional or no recommendation; the secondary outcome compared any recommendation (routine or conditional) with no recommendation. Reimbursement was classified as full, partial, or none. Full reimbursement was compared with partial or no reimbursement, while any reimbursement combined full and partial reimbursement. Among routinely or conditionally recommended vaccines, a recommendation–reimbursement gap was defined as the absence of full reimbursement. Missing and unclassifiable responses were excluded from the respective analyses.

Statistical analyses used chi-square test for group comparisons. Generalized linear mixed-effects models with a binomial distribution were fitted using maximum likelihood estimation with the Laplace approximation to assess differences in vaccine recommendation, reimbursement, and the recommendation–reimbursement gap by vaccine type and jurisdiction income group, accounting for clustering within jurisdictions. All models included vaccine type, income group, and their interaction as fixed effects, with a random intercept for jurisdiction. The slice p-value tests the overall difference among the three income groups separately within each vaccine. Where the slice test was significant, Tukey-Kramer-adjusted pairwise comparisons were used for interpretation. Probabilities were model-estimated values with 95% confidence intervals. For the recommendation-reimbursement gap analysis, each vaccine-jurisdiction observation represented one vaccine assessed within one participating jurisdiction. We included vaccine-jurisdiction observations for which a vaccine was routinely or conditionally recommended and both recommendation and reimbursement data were available. The final analysis included 708 observations from 122 of the 124 participating jurisdictions, as recommendation and reimbursement data were not available for all vaccine-jurisdiction combinations.

All analyses were performed using GraphPad and SAS version 9.4 software. All tests were two-sided, with p<0.05 considered statistically significant.

## Results

A total of 124 responses representing 120 countries were included, comparing 38 LIC/LMIC, 36 UMIC, and 50 HIC (Figure 1). The four additional responses represented one additional region in Bosnia and Herzegovina, two additional Canadian provinces, and Puerto Rico.

**Figure 1.**
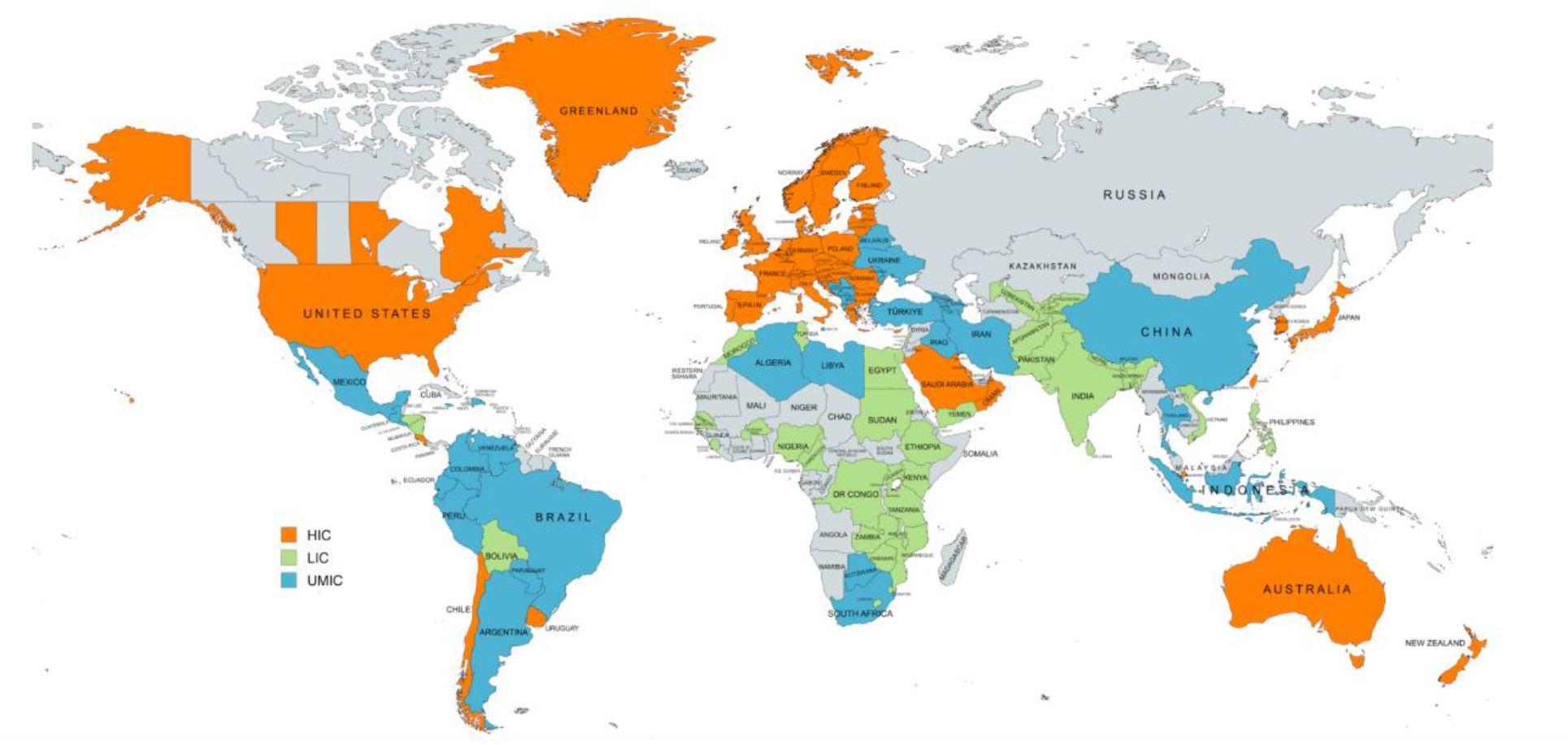
Geographic distribution of countries represented in the survey. A total of 124 responses represented 120 countries. Countries are color-coded according to the World Bank income classification: orange, high-income countries (HICs); blue, upper-middle-income countries (UMICs); and green, low- and lower-middle-income countries (LICs).

**Figure 2.**
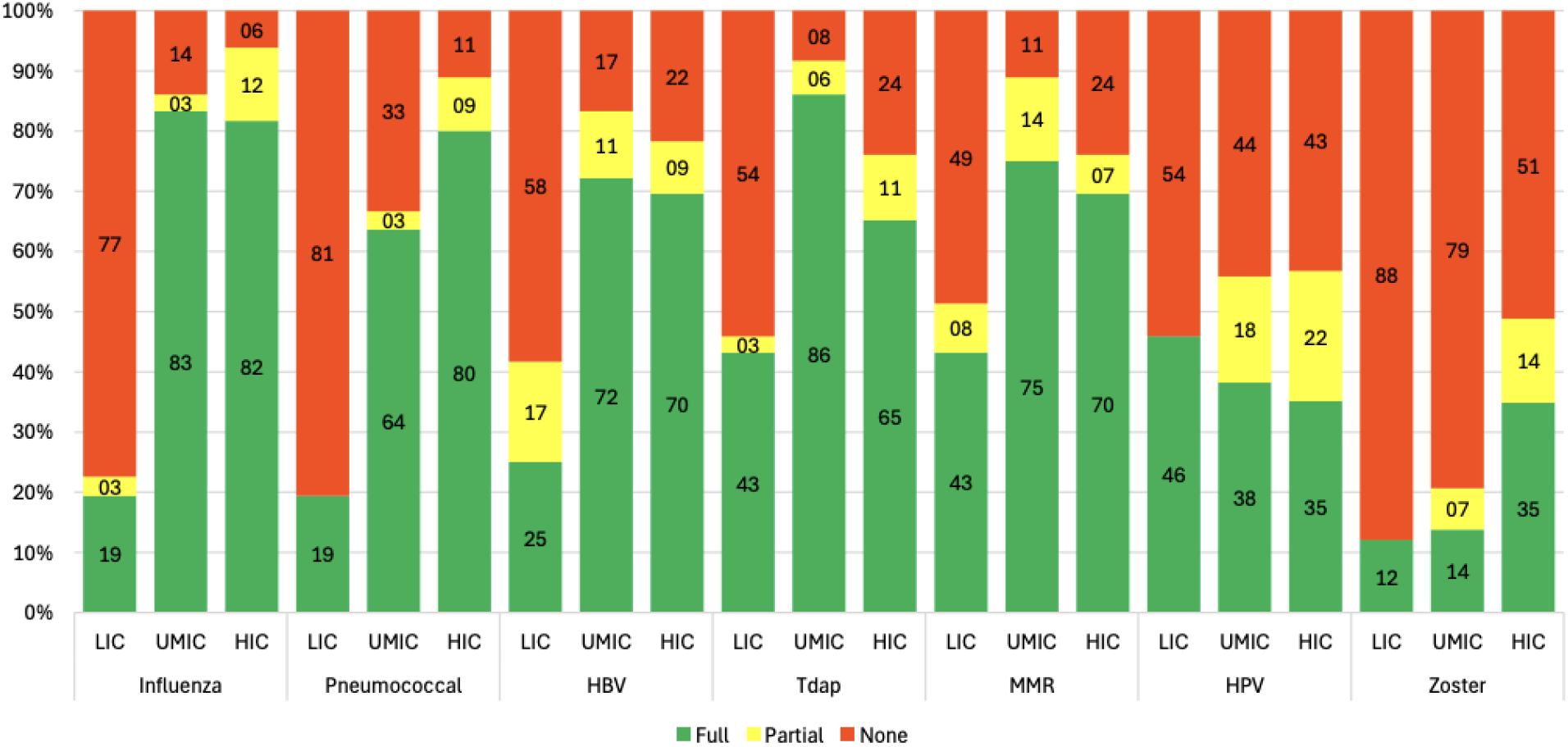
Public healthcare reimbursement of vaccines recommended for adults living with HIV, stratified by country income group. Vaccines are grouped by recommendation and presented according to country income classification. Green bars indicate full public reimbursement, yellow bars for partial reimbursement, and red bars with no public reimbursement.

Most respondents were infectious diseases physicians (69%), and over two-thirds had more than 10 years of experience providing HIV care (Table 1).

**Table 1.** Characteristics of survey respondents and participating countries (n=124 responses representing 120 countries)

| **Characteristic** | **n (%)** |
|---|---|
| **World Bank income classification** |  |
| LIC/LMIC | 38 (30.6%) |
| UMIC | 36 (29.0%) |
| HIC | 50 (40.3%) |
| **Respondent specialty** |  |
| Infectious diseases physician | 86 (69.4%) |
| General practitioner | 9 (7.3%) |
| Pediatrician | 3 (2.4%) |
| Other physician specialty | 14 (11.3%) |
| Registered nurse | 1 (0.8%) |
| Other healthcare professional | 11 (8.9%) |
| **Years of experience in HIV Care** |  |
| <5 years | 18 (14.5%) |
| 5-9 years | 22 (17.7%) |
| 10-20 years | 38 (30.6%) |
| >20 years | 46 (37.1%) |

Abbreviations: HIC, high-income country; HIV, human immunodeficiency virus; LIC, low- and lower-middle-income country; UMIC, upper-middle-income country.

### Vaccination guidelines and recommendations

HIV-specific national vaccination guidelines were reported by 46 of 124 respondents (37%) and availability differed significantly by country income classification from 18% in LIC/LMIC to 52% in HIC; p=0.026 (Table 2). General national vaccination recommendations were reported by 82 respondents (66%) and did not differ significantly across income groups. (Table 2). The use of EACS guidelines was reported by 61 respondents (49%) and increased significantly with income group (p<0.001), whereas the use of CDC recommendations was similar across income groups (Table 2).

**Table 2.** National vaccination guidance, reimbursement, and availability for people living with HIV.

| **Variable, n (%)** | **LIC/LMIC<br>(n=38)** | **UMIC<br>(n=36)** | **HIC<br>(n=50)** | ***p* value** |
|---|---|---|---|---|
| **National HIV-specific vaccination guidelines available** | 7 (18.4%) | 13 (36.1%) | 26 (52.0%) | 0.026 |
| **National health authority vaccination recommendations available** | 24 (63.2%) | 23 (63.9%) | 35 (70.0%) | 0.843 |
| **Use of EACS recommendations** | 8 (21.1%) | 22 (61.1%) | 31 (62.0%) | <0.001 |
| **Use of CDC recommendations** | 19 (50.0%) | 15 (41.7%) | 19 (38.0%) | 0.524 |
| **All recommended vaccines publicly reimbursed** | 5 (13.2%) | 10 (27.8%) | 22 (44.0%) | 0.007 |
| **Vaccination available within HIV clinic** | 21 (55.3%) | 25 (69.4%) | 39 (78.0%) | 0.02 |

Abbreviations: CDC, Centers for Disease Control and Prevention; EACS, European AIDS Clinical Society; HIC, high-income country; LIC/LMIC, low- and lower-middle-income country; UMIC, upper-middle-income country.
†Comparison of complete public reimbursement versus incomplete/no reimbursement.

Among the 852 evaluable vaccine-specific recommendation responses, 550 (64.6%) indicated routine recommendation, 203 (23.8%) conditional or selective recommendation, and 99 (11.6%) no recommendation or active promotion. In the mixed-effects analysis, routine recommendation differed significantly by vaccine type (p<0.0001) and income group (p=0.0010). The vaccine-by-income-group interaction was not significant (p=0.2979) and was not retained in the final model. The adjusted probability of routine recommendation increased from 52.6% (95% CI 41.2%-63.8%) in LIC/LMIC to 74.8% (95% CI 64.3%-83.1%) in UMIC and 77.6% (95% CI 69.1% %-84.3%) in HIC. After Tukey-Kramer adjustment, LIC/LMIC had lower odds of routine recommendation than UMIC (OR 0.37, adjusted p=0.0129) and HIC (OR 0.32, adjusted p=0.0013), while no difference was observed between UMIC and HIC (OR 0.86, adjusted p=0.8897).

Routine recommendation also differed by vaccine (Supplementary Table S2B). The adjusted probability was highest for pneumococcal vaccine (88.8%, 95% CI 81.4–93.4) and followed by HPV (83.8%, 95% CI 75.1–89.9) and lowest for zoster vaccine (42.6%) (Supplementary Figure 1). After correction for multiple comparisons, pneumococcal vaccination was significantly more likely to be routinely recommended than other vaccines except HPV, and zoster the least likely of all (Supplementary Table 2).

In the secondary analysis of any recommendation, defined as routine or conditional recommendation versus no recommendation, both vaccine type (p<0.0001) and income group (p=0.0012) were significant. The adjusted probability of any recommendation was high across all income groups: 90.2% in LIC/LMIC, 96.5% in UMIC, and 98.6% in HIC. Across vaccines, it ranged from 82.9% for zoster to 99.4% for hepatitis B. As several vaccine-income combinations had no non-recommendation responses, the interaction model produced unstable estimates. Therefore, the reduced main-effects model was retained and this outcome was treated as a secondary analysis.

### Vaccines reimbursement

Only 37 respondents (30%) reported that all recommended vaccines were fully reimbursed through the public healthcare system. Partial reimbursement was reported by 58 respondents (47%), while 25 (20%) reported that vaccines were paid entirely by patients. Full reimbursement was more common in countries with HIV-specific national vaccination guidelines than in those without (41.3% vs. 21.1%), although the difference was not statistically significant (p=0.133). Full reimbursement differed significantly by country income p=0.007), increasing across income groups (Table 2). Among the 799 evaluable vaccine-specific reimbursement responses, 425 (53.2%) indicated full reimbursement, 68 (8.5%) partial reimbursement, and 306 (38.3%) no reimbursement.

Complete reimbursement of all recommended vaccines was reported by 13% of LIC/LMIC, 28% of UMIC, and 44% of HIC.

In the mixed-effects analysis, full reimbursement differed significantly by vaccine type, income group, and the vaccine-by-income interaction (all p<0.0001); income-group effects were therefore evaluated separately within each vaccine (Figure 3). The largest income-related differences were observed for influenza, pneumococcal, and hepatitis B vaccination, where full reimbursement was very low in LIC/LMIC but near-universal in higher-income jurisdictions; for influenza, for example, the adjusted probability rose from approximately 3% in LIC/LMIC to about 97% in UMIC and HIC (Figure 3). After Tukey-Kramer correction, LIC/LMIC had lower odds of full reimbursement than both UMIC and HIC for influenza, pneumococcal, hepatitis B, and MMR vaccination. For Tdap, the difference between LIC/LMIC and UMIC remained significant, while comparisons involving HIC were no longer significant after adjustment.

**Figure 3.**
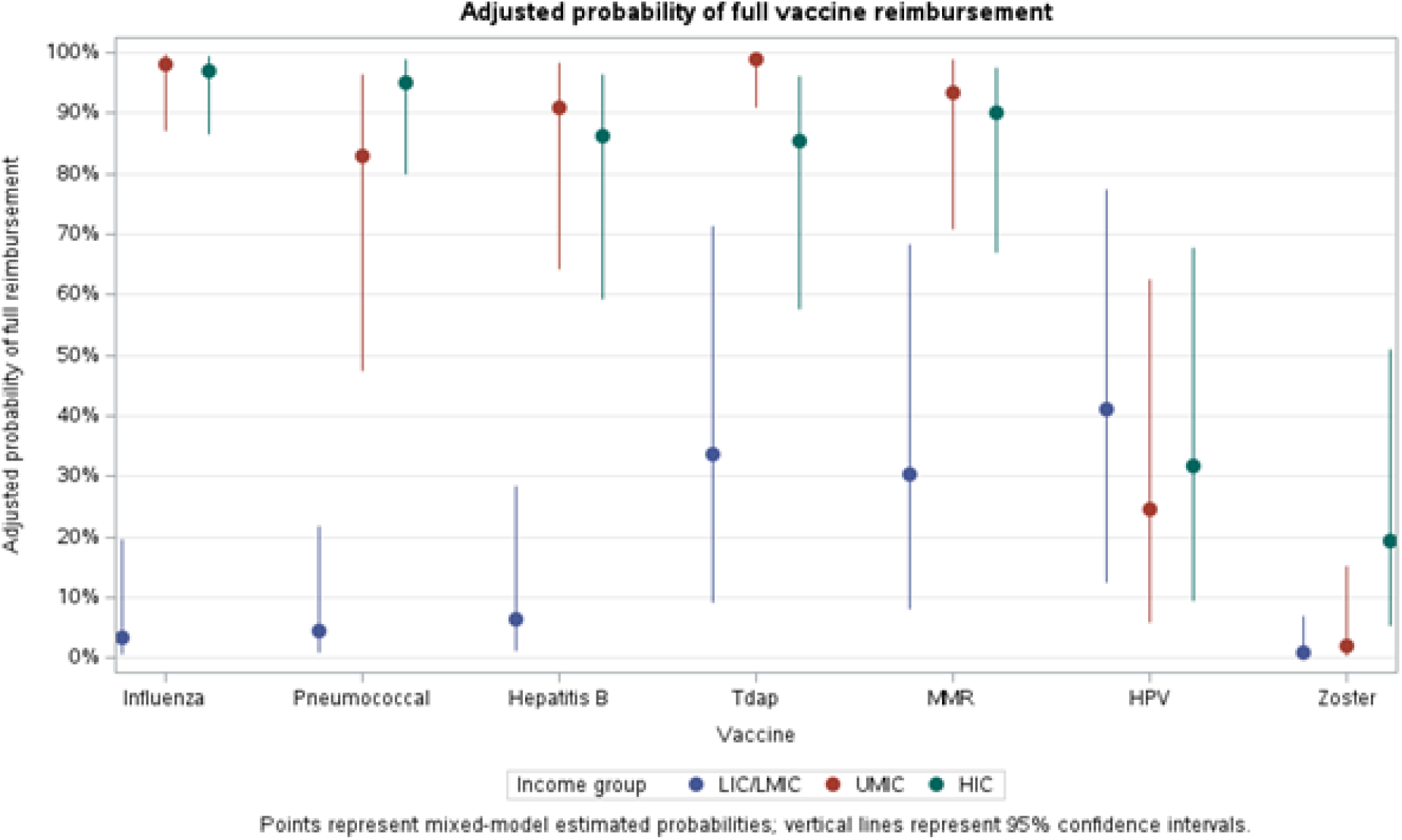
Adjusted probability of full vaccine reimbursement of a vaccine that is routinely or conditionally recommended by vaccine type and jurisdiction income group, with 95% confidence intervals.

Zoster had the lowest overall reimbursement among the vaccines evaluated. remaining uncommon across all income groups (from 12% LIC/LMIC to 35% HIC), including high-income settings (p=0.027; Table 3). Full reimbursement for HPV did not differ significantly by income group in the adjusted analysis (p=0.8012, Supplementary Table 3) and was, if anything, somewhat higher in lower-income settings (Table 3). Results for any reimbursement (full or partial versus none) were consistent with those for full reimbursement, with significant income-group differences for all vaccines except HPV (Supplementary Table 3).

**Table 3.** Percentage of full reimbursement for each vaccine by World Bank income classification.

| **Vaccine** | **LIC/LMIC** | **UMIC** | **HIC** | ***p*** |
|---|---|---|---|---|
| **Influenza** | 19.4% | 83.3% | 81.6% | <0.001 |
| **Pneumococcal** | 19.4% | 63.6% | 80.0% | <0.001 |
| **Hepatitis B** | 25.0% | 72.2% | 69.6% | <0.001 |
| **Tdap** | 43.2% | 86.1% | 65.2% | <0.001 |
| **MMR** | 43.2% | 75.0% | 69.6% | 0.0097 |
| **HPV** | 45.9% | 38.2% | 35.1% | 0.6205 |
| **Zoster** | 12.1% | 13.8% | 34.9% | 0.027 |

In the secondary analysis of any reimbursement, defined as full or partial reimbursement versus no reimbursement, vaccine type, income group, and their interaction were all significant (all p<0.0001). Income-group differences were significant for influenza, pneumococcal, hepatitis B, MMR, Tdap, and zoster vaccination, but not for HPV, similar to the findings for full reimbursement (Supplementary Table 3).

### Recommendation-reimbursement gap

The recommendation–reimbursement-gap analysis included 708 observations from 122 jurisdictions. The probability that a recommended vaccine was not fully reimbursed differed significantly by vaccine type (p<0.0001), income group (p=0.0013), and the vaccine-by-income-group interaction (p<0.0001). In LIC/LMIC jurisdictions, the adjusted probability of full reimbursement among recommended vaccines was only 4.3% for influenza, 4.1% for pneumococcal, and 5.4% for hepatitis B vaccination, compared with substantially higher probabilities in UMIC and HIC jurisdictions. (Figure 3); a similar but less pronounced pattern was seen for Tdap and MMR vaccination (Supplementary Tables S2A-S2D). For zoster vaccination, the gap was high across all income groups, as was the gap for HPV vaccination, persisting even in high-income settings. Income-group differences in the gap were statistically significant for influenza pneumococcal, hepatitis B, Tdap, and zoster vaccination, but not for HPV or MMR (Supplementary Tables S3A-S3D).

### Integration of vaccination into HIV care

Availability of vaccination directly within HIV clinics also differed significantly by income (p=0.020), increasing from 55% in LIC/LMIC to 78% in HIC jurisdictions (Table 2). Respondents from countries without on-site vaccination most reported system-level barriers, including vaccines being administered exclusively by general practitioners, insufficient staffing, and limited clinic resources.

## Discussion

In this global survey of 120 countries, vaccination was widely recommended for adults living with HIV, but substantial disparities remained in reimbursement and delivery. Routine recommendations were less frequent in LIC/LMIC, while reimbursement varied by both income group and vaccine type, highlighting a disconnect between vaccine recommendations and public financial support.

More than 90% of vaccines were estimated to be recommended either routinely or conditionally across all income groups, suggesting that vaccination is widely recognized as part of HIV care. However, routine recommendations remained significantly lower in LIC/LMIC than in UMIC and HIC. Influenza and pneumococcal vaccination are generally recommended for people living with HIV, whereas hepatitis B, MMR, and HPV recommendations may depend on previous immunity status, age, or previous vaccination [10-12]. This distinction is important as some vaccines are only recommended for certain patients, and this does not necessarily indice a gap in national vaccination recommendations.

Despite widespread national and international vaccination recommendations, public reimbursement remained limited, with only 30% of respondents reporting full reimbursement for all recommended vaccines. This highlights an important gap between recognizing the need for vaccination in adults living with HIV and providing the financial support needed to access it. Previous studies have mainly focused on patient- and provider-level barriers to vaccination among people living with HIV, including knowledge, vaccine hesitancy, healthcare-provider recommendation, and socioeconomic factors [13,14]. Our findings extend this research but identifying reimbursement policies as an additional barrier to vaccine access, as out of pocket costs may discourage uptake even when vaccination is recommended. [15,16]

The significant vaccine-by-income interaction for reimbursement showed that income-related differences varied considerably by vaccine. The largest differences were observed for influenza, pneumococcal, and hepatitis B vaccination, for which full reimbursement was very low in LIC/LMIC but substantially higher in UMIC and HIC. This is particularly concerning for commonly recommended vaccines, as lower reimbursement may place a greater financial burden on patients in countries with less resources. WHO has reported that adult vaccination remains underdeveloped in many lower-income countries, where immunization programs have historically focused on children, adolescents, and pregnant women [17]. However, income alone does not explain these differences, as reimbursement varied by vaccine and was not always available even in HIC [15].

HPV and zoster vaccination differed from the general reimbursement pattern. HPV was frequently routinely recommended, but reimbursement remained limited. Unlike most other vaccines, HPV reimbursement did not differ significantly by income group, suggesting that vaccine-specific policies and eligibility criteria may be important factors beyond national income. Differences in national HPV vaccination programs, particularly age restrictions and eligibility for publicly funded vaccination may help explain this pattern [18-20]. Zoster had the lowest overall reimbursement, including in HIC. This is particularly important because people living with HIV remain at increased risk of herpes zoster despite effective antiretroviral therapy [21], and recombinant zoster vaccination is recommended for people living with HIV [10]. Vaccine cost, age restrictions, and its more recent inclusion in vaccination programs may contribute to limited reimbursement [22,23].

Beyond reimbursement, our findings also identified differences in how vaccination is delivered within HIV care. Previous studies have shown that guideline availability does not necessarily translate into high vaccination coverage and that missed healthcare-provider recommendations remain an important barrier [8, 24-25]. In our study, vaccination was more frequently available directly within HIV clinics in HIC, while respondents without on-site vaccination commonly reported insufficient staffing and limited clinic resources. Integrating vaccination into routine HIV care may reduce some of these barriers by allowing vaccination during scheduled HIV visits and reducing the need for additional appointments [22, 26-27].

A major strength of this study is its broad international participation, representing 120 countries and evaluating factors affecting vaccine access, including reimbursement and service delivery. By comparing individual vaccines across income groups, we identified differences in reimbursement that may not be apparent when vaccination policies are considered. Several limitations should also be considered. Information for each country or jurisdiction was generally provided by a single expert and may not reflect the practices of other healthcare professionals or regional differences within national healthcare systems. Responses were self-reported and were not independently verified against national policy documents. Furthermore, harmonization of vaccine-specific responses into broader recommendation categories may have obscured differences between clinical indications and practical constraints affecting vaccination practices. The survey evaluated reimbursement and vaccine availability rather than patient-level vaccination coverage, so it was not possible to determine whether reported policies translated into actual vaccine uptake. In addition, our focus on public reimbursement may not capture vaccine access through donor-supported or non-governmental programs, particularly in lower-income settings. Finally, vaccination recommendations and reimbursement policies continue to evolve, and our findings reflect practices at the time the survey was conducted.

## Conclusion

Important disparities in vaccination access for adults living with HIV persist despite vaccination being widely recommended across diverse income groups. Public reimbursement showed greater differences than recommendations, varying by vaccine type and country income level. Moreover, we identified that commonly recommendations were not aligned with reimbursement policy, creating an important gap in implementation. Improving vaccine access will therefore require not only clear vaccination recommendations, but also adequate financing and practical strategies to integrate vaccination into routine HIV care.

## Data Availability

All data produced in the present study are available upon reasonable request to the authors.

## SUPPLEMENTARY MATERIALS

**Supplementary Material 1: Questionnaire**

**Supplementary Material 2: Supplementary Tables**

