## Supplementary Tables for "Global Disparities in Vaccine Reimbursement for Adults Living with HIV: A Cross-Sectional Survey of 120 Countries by EVASG and ESTISG"

**Supplementary Table S1. Detailed vaccine recommendations and reimbursement stratified by vaccine type and income group**

| SURVEY GROUP | | | VACCINE RECOMMENDATION | | | | PUBLIC REIMBURSEMENT | | | |
|---|---|---|---|---|---|---|---|---|---|---|
| **Vaccine** | **Income group** | **Jurisdictions, N** | **Responses, N** | **Routine, n (%)** | **Conditional, n (%)** | **Not recommended, n (%)** | **Responses, N** | **Fully reimbursed, n (%)** | **Partially reimbursed, n (%)** | **Not reimbursed, n (%)** |
| **Influenza** | **Overall** | **124** | **118** | **78 (66.1%)** | **28 (23.7%)** | **12 (10.2%)** | **116** | **76 (65.5%)** | **8 (6.9%)** | **32 (27.6%)** |
| | LIC/LMIC | 38 | 35 | 14 (40.0%) | 11 (31.4%) | 10 (28.6%) | 31 | 6 (19.4%) | 1 (3.2%) | 24 (77.4%) |
| | UMIC | 36 | 35 | 26 (74.3%) | 9 (25.7%) | 0 (0.0%) | 36 | 30 (83.3%) | 1 (2.8%) | 5 (13.9%) |
| | HIC | 50 | 48 | 38 (79.2%) | 8 (16.7%) | 2 (4.2%) | 49 | 40 (81.6%) | 6 (12.2%) | 3 (6.1%) |
| **Pneumococcal** | **Overall** | **124** | **123** | **103 (83.7%)** | **11 (8.9%)** | **9 (7.3%)** | **114** | **64 (56.1%)** | **5 (4.4%)** | **45 (39.5%)** |
| | LIC/LMIC | 38 | 38 | 28 (73.7%) | 5 (13.2%) | 5 (13.2%) | 36 | 7 (19.4%) | 0 (0.0%) | 29 (80.6%) |
| | UMIC | 36 | 36 | 29 (80.6%) | 3 (8.3%) | 4 (11.1%) | 33 | 21 (63.6%) | 1 (3.0%) | 11 (33.3%) |
| | HIC | 50 | 49 | 46 (93.9%) | 3 (6.1%) | 0 (0.0%) | 45 | 36 (80.0%) | 4 (8.9%) | 5 (11.1%) |
| **Hepatitis B** | **Overall** | **124** | **122** | **66 (54.1%)** | **53 (43.4%)** | **3 (2.5%)** | **118** | **67 (56.8%)** | **14 (11.9%)** | **37 (31.4%)** |
| | LIC/LMIC | 38 | 38 | 16 (42.1%) | 19 (50.0%) | 3 (7.9%) | 36 | 9 (25.0%) | 6 (16.7%) | 21 (58.3%) |
| | UMIC | 36 | 36 | 24 (66.7%) | 12 (33.3%) | 0 (0.0%) | 36 | 26 (72.2%) | 4 (11.1%) | 6 (16.7%) |
| | HIC | 50 | 48 | 26 (54.2%) | 22 (45.8%) | 0 (0.0%) | 46 | 32 (69.6%) | 4 (8.7%) | 10 (21.7%) |
| **Tdap** | **Overall** | **124** | **123** | **84 (68.3%)** | **24 (19.5%)** | **15 (12.2%)** | **119** | **77 (64.7%)** | **8 (6.7%)** | **34 (28.6%)** |
| | LIC/LMIC | 38 | 38 | 21 (55.3%) | 11 (28.9%) | 6 (15.8%) | 37 | 16 (43.2%) | 1 (2.7%) | 20 (54.1%) |
| | UMIC | 36 | 36 | 26 (72.2%) | 6 (16.7%) | 4 (11.1%) | 36 | 31 (86.1%) | 2 (5.6%) | 3 (8.3%) |
| | HIC | 50 | 49 | 37 (75.5%) | 7 (14.3%) | 5 (10.2%) | 46 | 30 (65.2%) | 5 (10.9%) | 11 (23.9%) |
| **MMR** | **Overall** | **124** | **122** | **68 (55.7%)** | **34 (27.9%)** | **20 (16.4%)** | **119** | **75 (63.0%)** | **11 (9.2%)** | **33 (27.7%)** |
| | LIC/LMIC | 38 | 38 | 18 (47.4%) | 13 (34.2%) | 7 (18.4%) | 37 | 16 (43.2%) | 3 (8.1%) | 18 (48.6%) |
| | UMIC | 36 | 35 | 19 (54.3%) | 11 (31.4%) | 5 (14.3%) | 36 | 27 (75.0%) | 5 (13.9%) | 4 (11.1%) |
| | HIC | 50 | 49 | 31 (63.3%) | 10 (20.4%) | 8 (16.3%) | 46 | 32 (69.6%) | 3 (6.5%) | 11 (23.9%) |
| **HPV** | **Overall** | **124** | **122** | **96 (78.7%)** | **18 (14.8%)** | **8 (6.6%)** | **108** | **43 (39.8%)** | **14 (13.0%)** | **51 (47.2%)** |
| | LIC/LMIC | 38 | 38 | 27 (71.1%) | 7 (18.4%) | 4 (10.5%) | 37 | 17 (45.9%) | 0 (0.0%) | 20 (54.1%) |
| | UMIC | 36 | 35 | 31 (88.6%) | 3 (8.6%) | 1 (2.9%) | 34 | 13 (38.2%) | 6 (17.6%) | 15 (44.1%) |

| SURVEY GROUP | | | VACCINE RECOMMENDATION | | | | PUBLIC REIMBURSEMENT | | | |
|---|---|---|---|---|---|---|---|---|---|---|
| **Vaccine** | **Income group** | **Jurisdictions, N** | **Responses, N** | **Routine, n (%)** | **Conditional, n (%)** | **Not recommended, n (%)** | **Responses, N** | **Fully reimbursed, n (%)** | **Partially reimbursed, n (%)** | **Not reimbursed, n (%)** |
| | HIC | 50 | 49 | 38 (77.6%) | 8 (16.3%) | 3 (6.1%) | 37 | 13 (35.1%) | 8 (21.6%) | 16 (43.2%) |
| **Zoster** | **Overall** | **124** | **122** | **55 (45.1%)** | **35 (28.7%)** | **32 (26.2%)** | **105** | **23 (21.9%)** | **8 (7.6%)** | **74 (70.5%)** |
| | LIC/LMIC | 38 | 38 | 12 (31.6%) | 8 (21.1%) | 18 (47.4%) | 33 | 4 (12.1%) | 0 (0.0%) | 29 (87.9%) |
| | UMIC | 36 | 35 | 16 (45.7%) | 9 (25.7%) | 10 (28.6%) | 29 | 4 (13.8%) | 2 (6.9%) | 23 (79.3%) |
| | HIC | 50 | 49 | 27 (55.1%) | 18 (36.7%) | 4 (8.2%) | 43 | 15 (34.9%) | 6 (14.0%) | 22 (51.2%) |

**Note**. Values are n (%) among non-missing responses; recommendation and reimbursement denominators may differ.

**S2. Vaccine recommendation models**

**Supplementary Table S2A. Overall mixed-effects logistic model results for routine and any recommendation**

| **Outcome** | **Observations** | **Jurisdictions** | **Vaccine effect, p** | **Income-group effect, p** | **Vaccine–income interaction** | **Random-intercept variance** |
|---|---|---|---|---|---|---|
| Routine recommendation | 852 | 123 | <0.0001 | 0.001 | Not retained; initial p = 0.2979 | 1.2961 |
| Any recommendation | 852 | 123 | <0.0001 | 0.0012 | Not retained because of sparse cells and separation | 2.9053 |

**Supplementary Table S2B. Model-estimated recommendation probabilities by vaccine**

| Vaccine | Routine recommendation, % (95% CI) | Any recommendation, % (95% CI) |
|---|---|---|
| **Influenza** | 70.0 (58.9–79.2) | 96.0 (90.8–98.3) |
| **Pneumococcal** | 88.8 (81.4–93.4) | 97.6 (93.8–99.1) |
| **Hepatitis B** | 54.6 (43.1–65.6) | 99.4 (97.5–99.9) |
| **Tdap** | 72.5 (61.9–81.1) | 95.0 (89.2–97.8) |
| **MMR** | 56.7 (45.2–67.5) | 92.1 (84.4–96.1) |
| **HPV** | 83.8 (75.1–89.9) | 97.9 (94.4–99.2) |
| **Zoster** | 42.6 (31.8–54.0) | 82.9 (71.6–90.3) |

**Supplementary Table S2C. Model-estimated recommendation probabilities by income group**

| Income group | Routine recommendation, % (95% CI) | Any recommendation, % (95% CI) |
|---|---|---|
| **LIC/LMIC** | 52.6 (41.2–63.8) | 90.2 (81.2–95.1) |
| **UMIC** | 74.8 (64.3–83.1) | 96.5 (91.9–98.6) |
| **HIC** | 77.6 (69.1–84.3) | 98.6 (96.2–99.5) |

**Supplementary Table S2D. Tukey-adjusted income-group comparisons for routine recommendation**

| Comparison | Odds ratio | Tukey-adjusted 95% CI | Adjusted p-value |
|---|---|---|---|
| **LIC/LMIC versus UMIC** | 0.374 | 0.166–0.844 | 0.0129 |
| **LIC/LMIC versus HIC** | 0.321 | 0.150–0.685 | 0.0013 |
| **UMIC versus HIC** | 0.858 | 0.393–1.872 | 0.8897 |

### S3. Reimbursement and policy-gap models

**Supplementary Table S3A. Overall mixed-effects logistic model results for reimbursement and the recommendation–reimbursement gap**

| Outcome | Observations | Jurisdictions | Vaccine effect, p | Income-group effect, p | Vaccine × income interaction, p | Random-intercept variance |
|---|---|---|---|---|---|---|
| **Full reimbursement** | 799 | 124 | <0.0001 | <0.0001 | <0.0001 | 11.7812 |
| **Any reimbursement** | 799 | 124 | <0.0001 | <0.0001 | <0.0001 | 10.5108 |
| **Recommendation–reimbursement gap** | 708 | 122 | <0.0001 | 0.0013 | <0.0001 | 16.5529 |

**Note**. Effects and interactions are from mixed-effects logistic models. The recommendation–reimbursement gap is evaluated among vaccines classified as routinely or conditionally recommended.

**Supplementary Table S3B. Model-estimated probability of full reimbursement**

| Vaccine | LIC/LMIC, % (95% CI) | UMIC, % (95% CI) | HIC, % (95% CI) | Income-group slice p-value |
|---|---|---|---|---|
| **Influenza** | 3.4 (0.5–19.5) | 98.0 (87.1–99.7) | 96.9 (86.6–99.4) | <0.0001 |
| **Pneumococcal** | 4.4 (0.7–21.9) | 83.0 (47.4–96.3) | 95.0 (80.0–98.9) | <0.0001 |
| **Hepatitis B** | 6.5 (1.2–28.5) | 90.9 (64.2–98.2) | 86.3 (59.3–96.5) | 0.0001 |
| **Tdap** | 33.5 (9.2–71.4) | 98.8 (91.0–99.8) | 85.3 (57.7–96.1) | 0.001 |
| **MMR** | 30.4 (8.1–68.3) | 93.4 (70.7–98.8) | 90.0 (67.1–97.6) | 0.0076 |
| **HPV** | 41.2 (12.5–77.4) | 24.5 (6.0–62.4) | 31.8 (9.4–67.7) | 0.8012 |
| **Zoster** | 0.8 (0.1–6.8) | 2.0 (0.2–15.2) | 19.3 (5.2–51.0) | 0.0247 |

**Supplementary Table S3C. Model-estimated probability of any reimbursement**

| Vaccine | LIC/LMIC, % (95% CI) | UMIC, % (95% CI) | HIC, % (95% CI) | Income-group slice p-value |
|---|---|---|---|---|

| | | | | |
|---|---|---|---|---|
| **Influenza** | 7.0 (1.4–29.0) | 98.5 (89.6–99.8) | 99.7 (97.5–100.0) | <0.0001 |
| **Pneumococcal** | 6.1 (1.2–25.3) | 85.5 (52.5–96.9) | 98.5 (91.3–99.8) | <0.0001 |
| **Hepatitis B** | 30.3 (8.9–65.9) | 97.6 (85.7–99.6) | 94.2 (77.3–98.7) | 0.0005 |
| **Tdap** | 38.8 (12.6–73.6) | 99.6 (95.5–100.0) | 94.0 (77.1–98.7) | 0.0002 |
| **MMR** | 47.3 (17.1–79.7) | 99.1 (92.9–99.9) | 93.9 (76.7–98.7) | 0.0011 |
| **HPV** | 40.4 (13.4–74.7) | 66.1 (28.2–90.6) | 78.0 (44.2–94.0) | 0.3067 |
| **Zoster** | 1.3 (0.2–9.5) | 4.4 (0.7–23.7) | 47.2 (18.1–78.2) | 0.0022 |

**Supplementary Table S3D. Model-estimated probability of a recommendation–reimbursement gap**

| Vaccine | LIC/LMIC, % (95% CI) | UMIC, % (95% CI) | HIC, % (95% CI) | Income-group slice p-value |
|---|---|---|---|---|
| **Influenza** | 95.7 (67.5–99.6) | 0.9 (0.1–9.0) | 1.3 (0.2–8.7) | <0.0001 |
| **Pneumococcal** | 95.9 (72.8–99.5) | 8.1 (1.1–41.2) | 2.7 (0.4–15.3) | <0.0001 |
| **Hepatitis B** | 94.6 (69.0–99.3) | 5.3 (0.7–31.2) | 10.4 (2.0–39.9) | 0.0004 |
| **Tdap** | 71.3 (25.5–94.7) | 0.9 (0.1–9.7) | 10.5 (2.0–40.0) | 0.0032 |
| **MMR** | 53.6 (13.7–89.4) | 3.2 (0.3–23.9) | 8.8 (1.6–36.6) | 0.0611 |
| **HPV** | 56.0 (15.7–89.8) | 81.5 (38.1–96.9) | 66.0 (25.0–91.9) | 0.6646 |
| **Zoster** | 98.8 (85.7–99.9) | 99.3 (89.5–100.0) | 72.1 (31.6–93.5) | 0.024 |

**Note**. Values are model-estimated percentages with 95% CI. A gap denotes partial or no reimbursement among recommended vaccines; the final column gives the income-group slice p-value.
